# Cognitive Trajectories of Post-Stroke Delirium: Longitudinal Global and Domain-Specific Impairment and Digital Markers

**DOI:** 10.64898/2026.10.21.26366183

**Authors:** Lena S. Geiger, Franz Wortha, Sebastian Klütz, Christian Mychajliw, Christoph Gäbele, Andreas Jooß, Sophia Kindzierski, Jennifer Sartor-Pfeiffer, Constanze Single, Ulf Ziemann, Annerose Mengel

## Abstract

**Background:** Poststroke cognitive impairment (PSCI) and poststroke delirium (PSD) remain underexplored. Longitudinal assessments of cognition before PSD onset and at follow-up are scarce, leaving the association between PSD and subsequent global and domain-specific PSCI unclear. Digital assessments using task-derived kinematic parameters may provide complementary insights into cognitive vulnerability and PSD risk.

**Methods:** In this prospective longitudinal study, patients with acute stroke underwent multidomain tablet-based neuropsychological assessment within the acute phase, before PSD onset, and at 90 days post-stroke. Global PSCI (gPSCI) and domain-specific PSCI (attention and processing speed, executive functions, verbal memory, language, and visuospatial-construction) were assessed using norm-referenced cognitive performance. Cognitive outcomes were compared between patients with and without PSD at both time points. Functional outcome, mortality, and quality of life were assessed at 90 days. Exploratory random forest analyses evaluated the discriminatory ability of acute-phase task-derived kinematic parameters.

**Results:** Among 31 patients with stroke, 10 developed PSD, 18 served as controls, and 3 dropped out. At 90 days, patients with PSD had more severe gPSCI than controls (*U*=19.500, *p*<0.001), whereas the acute-phase difference was not significant (U=50.500, *p*=0.057). Domain-specific analyses revealed distinct temporal patterns of cognitive impairment. During the acute phase, patients with subsequent PSD, showed significantly more severe impairments in attention and processing speed (U=29.500; *p*=0.003) and visuospatial-construction (U=31.500; *p*=0.004). At 90 days, differences in attention and processing speed persisted (*U*=21.000, *p* <0.001) with additional significant differences in executive functions (*U*=38.500, *p*=0.012) and language (*U*=45.500, *p*=0.031). Kinematic parameters showed discriminatory performance comparable to gPSCI (78.89% versus 81.50%). At 90 days, PSD was also associated with worse functional outcome (U=31.500, *p*=0.004), higher mortality (*p*=0.037), and lower mental quality of life (*p*=0.021).

**Conclusions:** PSD is associated with more severe global PSCI at 90 days post-stroke and distinct temporal pattern of domain-specific PSCI. Task-derived kinematic parameters may complement conventional cognitive scores. Larger multicenter studies are needed to validate these findings.

**Trial Registration:** ClinicalTrials.gov: NCT06680336. https://clinicaltrials.gov/study/NCT06680336

## 1. Introduction

Post-stroke delirium (PSD) is a common complication of acute stroke, affecting up to 40% of older patients ^1–3^. Its etiology is multifactorial, with advanced age, comorbidities, infections, pre-existing cognitive impairment and stroke severity among the major risk factors ^1, 4, 5^. PSD is associated with longer hospitalization, increased mortality, and lower quality of life ^1, 6–13^. Further, previous studies have reported poorer cognitive outcomes following delirium, particularly in global cognition and executive functions, compared with healthy controls ^14–17^. However, studies specifically examining cognitive outcomes after PSD remain scarce, and evidence regarding its association with subsequent cognitive decline is limited. Most studies evaluate cognition weeks to months after stroke ^18–20^ and rely on global screening scores or dichotomous definitions of post-stroke cognitive impairment (PSCI) ^13, 17, 19, 21^, potentially overlooking domain-specific deficits and heterogeneous cognitive trajectories ^22, 23^ ^24^. Early multidomain cognitive assessment may therefore enable a more detailed characterization of cognitive vulnerability and subsequent decline while providing insights into the specific contribution of PSD to the development and severity of PSCI. Digital assessments may additionally facilitate bedside testing and task-derived kinematic parameters may capture distinct behavioral patterns beyond conventional cognitive scores ^25, 26^.

To address this gap, we examined PSCI in a critically ill stroke population at two timepoints: first during the early acute phase within 48 hours after admission (≤ 72h stroke onset) and prior to the occurrence of PSD and at 90 days post-stroke, using a multidomain neuropsychological assessment partially incorporated within a digital application.

The primary endpoint was a between-group comparison in cognitive outcome, defined as global PSCI, at 90 days post-stroke. PSCI was characterized as either average cognitive functioning or mild, moderate or severe cognitive impairment, according to the performance level (norm-referenced z-scores) on a multidomain neuropsychological assessment. Secondary endpoints were between-group differences in global PSCI during the acute phase and the temporal pattern of domain-specific PSCI (attention and information speed, executive functions, verbal memory, language, visuospatial-construction). Additional secondary endpoints included between-group differences in mRS, NIHSS, mortality, quality of life and cognitive recovery at 90 days post-stroke. Further as an exploratory analysis, a machine learning approach using a random forest classifier and acute-phase task-derived kinematic parameters were evaluated for discriminatory ability between groups.

## 2. Methods

### 2.1. Study population & inclusion criteria

In this prospective longitudinal study, 31 patients (30 Caucasians, 1 Black) with an acute ischemic stroke were recruited between May 2024 and October 2025 from the stroke unit ward of the Neurological Department of the University Hospital Tübingen. Ischemic stroke was defined as focal onset of neurological symptoms lasting >24 hours and a National Institutes of Health Stroke Scale (NIHSS) ^27^ score of > 1. Stroke was confirmed by an CT scan on admission. Further inclusion criteria were age over 60 years, stroke onset within 96 hrs of enrolment, Glasgow coma scale (GCS) > 8. Exclusion criteria were delirium onset before study enrolment, global aphasia, pre-existing conditions such as seizures, traumatic head injury within past three month or pre-existing neuropsychiatric disorder e.g. dementia or developmental delay. A detailed description has been previously published ^28^.

### 2.2. Ethics approval

The study has been reviewed and approved by the local Ethics Committee of the Medical Faculty and the University Hospital Tuebingen (Ref.no. 444/2023BO2) and ethical principles for medical research involving human subjects of the Declaration of Helsinki, adopted by the 18th General Assembly of the World Medical Association (World Medical Association, 1964). Informed consent was obtained from all participants, or from legal guardians in cases where patients were unable to provide consent themselves. Testing was stopped immediately if signs of distress or fatigue were observed. Clinical staff remained available throughout all procedures.

### 2.3. Study procedure and Assessments

The first assessment (session S1) was conducted during the early acute phase post-stroke (≤ 48h after hospital admission, ≤ 72h after stroke onset and before PSD development) in an ICU-setting. The second session (S2) took place 90 days post-stroke as an outpatient assessment in hospital or home-based assessment (when patients’ condition or transfer precludes in-hospital assessment). See Figure 1 for more details.

**Figure 1.**
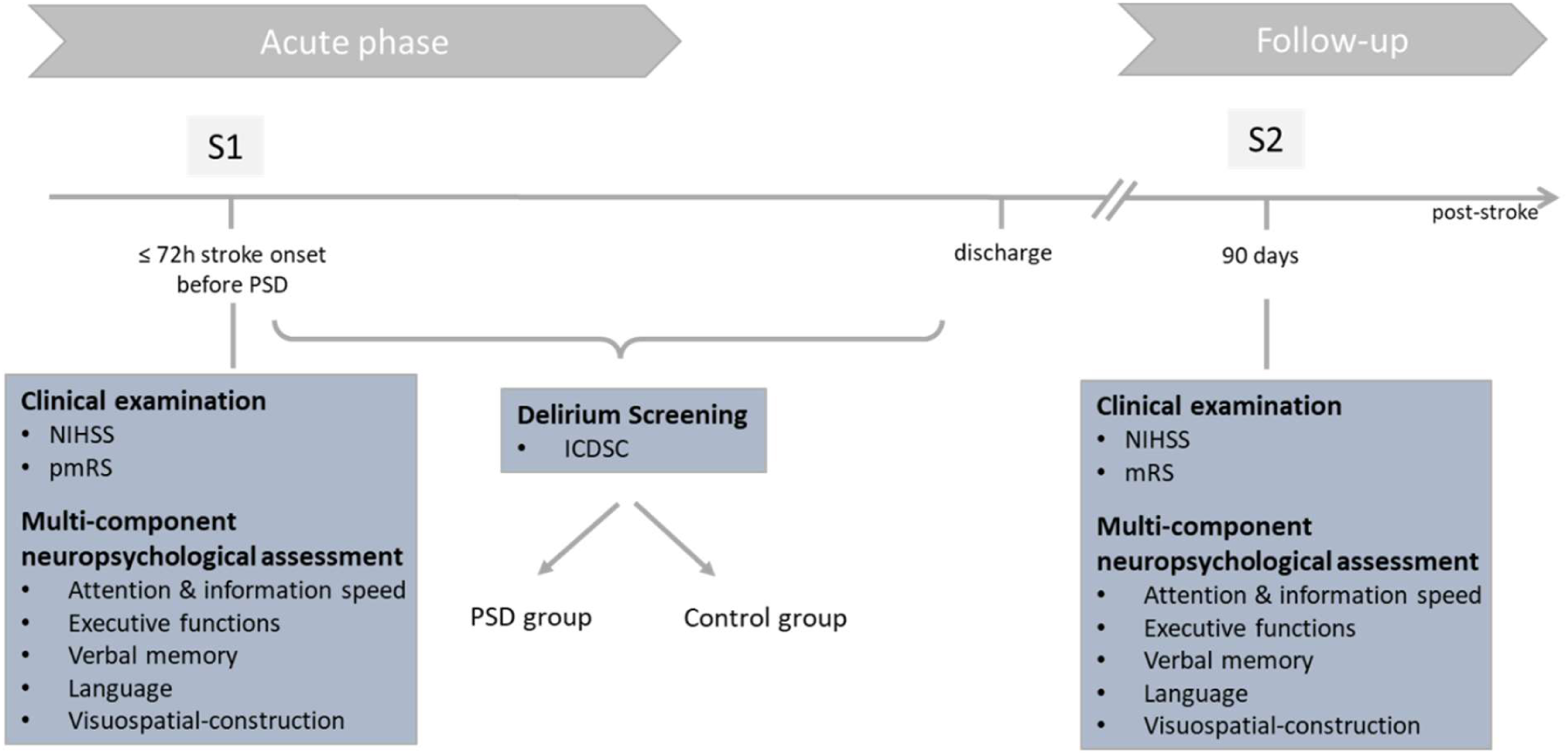
Overview of the RECOVER study procedures. Abbreviations: NIHSS = National Institutes of Health Stroke Scale, pmRS = pre-modified Rankin Score, ICDSC = Intensive Care Delirium Screening Checklist.

Each assessment (S1 – S2) took 45-60 minutes and consisted of a clinical examination (via NIHSS and modified Ranking scale (mRS)^29^ and a multi-component neuropsychological testing including the cognitive domains: attention and information speed, executive functions, verbal memory, language, visuospatial-construction and orientation (see Table 1). In general, questionnaires and clinical scores used were appropriate for our elderly study population, with available normative data and the selection of tests and domains aligns with the recommendations of major stroke organizations ^30, 31^. To reduce practice effects across sessions, we use official parallel test forms when available. A more detailed description of the neuropsychological assessment and clinical scores is provided in Table *1*. All drawing parts of the assessment (Trail making test, clock drawing, figure copying), as well as some questionnaires were digitalized using a tablet app called TuCAN (Tübingen Cognitive Assessment for Neuropsychiatric Disorders ^32^ and a digitizer-pencil, which has been designed for elderly individuals and has previously been evaluated in healthy and patient cohorts (stroke, cognitive impairment) ^25, 33–35^. To address potential unfamiliarity with digital technology in older stroke patients, patients completed a short familiarization task before the formal assessment.

**Table 1.** Cognitive Domains and subtests within the neuropsychological assessment.

| COGNITIVE DOMAINS | SUBTEST OR SUBSCALE |
| --- | --- |
| Attention & processing Speed | • Trail making test (TMT) – Part A <sup>36</sup><br>• NAB Module Attention <sup>37</sup> |
| Executive functions | • WM span (NAB Module Attention <sup>37</sup>)<br>• TMT – Part B <sup>36</sup> |
| Verbal memory | Memory index score (MIS) within MoCA <sup>38</sup> |
| Language | Language index score (LIS) within MoCA <sup>38, 39</sup> |
| Visuospatial-construction | Visuospatial index score (VIS) within MoCA <sup>38, 39</sup> |

Abbreviations: NAB = Neuropsychological Assessment Battery, WM = working memory, TMT = Trail Making Test.

Pre-stroke cognitive functioning was assessed by the relative questionnaire Informant Questionnaire on Cognitive Decline in the Elderly (IQCODE) ^40^ and the cognitive failure questionnaire ^41^. Quality of life was assessed using the SF-12 ^42^, further a depression assessment using the geriatric depression scale (GDS) ^43^ was administered. Some tests were derived from the Montreal Cognitive Assessment Test (MoCA)^38^. Further, demographic information and clinical data regarding stroke location, comorbidities or treatment have been assessed. Delirium screening, using the Intensive Care Delirium Screening Checklist (ICDSC) ^44^, was conducted throughout ICU treatment and was performed every 8 hours. Diagnosis was made according to DSM-5 criteria by an experienced neurologist who was blinded towards the behavioral outcome data. Delirium recovery was defined as three consecutive negative ICDSC screenings.

### Statistical analysis

#### Multi-component neuropsychological assessment

In general, analysis will be conducted using SPSS Statistics (Version 28.0.0.0, IBM Corp. Released 2023)^45^ and MATLAB (R2024a) ^46^.

In a first step, the analysis of the above-mentioned tests, questionnaires and clinical scores was conducted according to the applicable manual derived from the test publisher. Raw scores were transformed to age-, sex- and education-corrected *z* scores available from normative data or previous publications ^47, 48^.

In second step, a global and a domain-specific PSCI scores were defined in accordance with the International Society for Vascular Behavioral and Cognitive Disorders (VASCOG) ^49^ and similarly to previous studies ^20, 30, 50–52^. Further individual performance was defined as average cognitive functioning and mild, moderate or severe cognitive impairment according to the performance level on norm-referenced z-scores, which is in line with the recommendations of multiple neuropsychological societies ^53^. If a cognitive domain was represented by more than one tests, the mean of the z-scores was used to represent the cognitive domain. The following domains were defined: attention and information speed, executive functions, verbal memory, language, visuospatial-construction and orientation (see Table 1 for more details).

In detail, domain-specific (ds) PSCI was defined as a 5-level performance score. A *z* score ≤ - 1, was defined as average cognitive functioning (PSCI = 0). A *z* score ≥ -1 & ≤ - 2 (between 3rd and 16th percentile) was defined as mild cognitive impairment (PSCI = 1), *z* score ≥ -2 & ≤ - 3 (lower than 3rd percentile) was defined as moderate (PSCI = 2), while *z* score ≥ 3 (lower than 1st percentile) was defined as severe cognitive impairment (PSCI = 3). Due to increased mortality in the PSD group, and to accommodate clinical realities as well as access feasibility of our study protocol, we use an intention-to-treat analysis for the primary endpoint. Therefore, we have added a level (PSCI = 4) with reflects the death of a patient before the 3-month-follow up session. Global PSCI (gPSCI) was defined according to the level of performance in the five cognitive domains. A dsPSCI = 1 on at least one cognitive domain was defined as mild cognitive impairment (gPSCI = 1). A dsPSCI = 2 in at least two domains was defined as moderate (gPSCI = 2), while a dsPSCI = 3 in at least two cognitive domains or dsPSCI = 2 in most of the domains was defined as severe cognitive impairment (PSCI = 3). To minimize the potential impact of extreme scores on calculations, *z* scores were truncated at a minimum of *z* =−3 and a maximum of *z* =3. For missing data in a two-test subdomain, the *z* score of the other test within the same domain has been used.

For group differences, independent t-tests were used to compare continuous variables between groups, Mann-Whitney-*U* tests were used to compare ordinal variables between groups and χ^2^ test or Fisher’s exact tests were used to compare categorical variables. For all tests, we used a 2-sided significance level of 0.05. An exploratory ordinal logistic regression was performed due to the small sample size to estimate the association between post-stroke delirium and functional outcome (mRS), reporting odds ratios (ORs) with 95% confidence intervals (CIs) and p-values. The proportional odds assumption was assessed using the test of parallel lines.

The primary endpoint was defined a between-group comparison of cognitive outcome, defined as gPSCI (score 0 = no gPSCI, 4 = death) at 3-months follow-up (90 days). Secondary endpoints were between-group differences in global PSCI during the acute phase and the temporal pattern of domain-specific PSCI (attention & information speed, executive functions, verbal memory, language, visuospatial-construction; score 0 = no dsPSCI, 4 = death). Additional secondary endpoints included between group-differences in modified Rankin Scale (mRS; score 0 = no symptoms, 6 = death), mortality, good outcome (mRS 0-2), favourable outcome (mRS ≤ 3), NIHSS, quality of life (SF-12) and cognitive recovery at 90 days post-stroke. Further, digital parameters such as time in air or time on surface ^25, 26^ were used to evaluate their prognostic value for identifying patients with subsequent development of PSD. We focused on kinematic drawing features derived from the figure copying task within the MoCA which assess vvisuospatial-construction and executive functions and have previously been described as reflecting persisting impairments after stroke ^16, 54, 55^. For this exploratory analysis, a machine learning approach i.e., a random forest classifier was utilized within a 10-time repeated leave-one-out cross-validation (LOOCV) framework. To assess the relative utility of the drawing variables, models utilizing three distinct feature sets were fitted for each drawing task: (1) a control model utilizing the gPSCI during the acute phase post-stroke (S1), (2) a drawing features model incorporating time on surface, time above surface, the proportion between these two, and average on-surface velocity, and (3) a combined model incorporating both clinical and drawing features. Because the primary objective of this analysis was to examine how the different feature sets affect predictive capability rather than to maximize absolute predictive performance, hyperparameters were held fixed (e.g., maximum tree depth = 5) instead of nested hyperparameter optimization. To account for the stochastic variance of the random forest algorithm, patient-level classifications were aggregated via majority voting across the ten LOOCV repetitions. This generated a single, stable consensus prediction per patient for each of the three feature sets. Statistical comparison of model performance was then conducted using these consensus predictions. Cochran’s Q test was employed as an omnibus test to determine whether model performance significantly differed depending on the feature set. Where applicable, post-hoc pairwise McNemar’s tests were run with a Bonferroni correction to identify specific differences between models.

## 3. Results

### 3.1. Patient characteristics

A total of 31 acute stroke patients (16 males, 15 females) were enrolled from inpatient submissions. Three patients were excluded before the first assessment due to consent withdrawal (n = 2) or global aphasia following clinical deterioration leaving 28 patients for analysis (see Figure 2). Of these, ten patients (36%) subsequently developed PSD and 18 (64%) did not.

**Figure 2.**
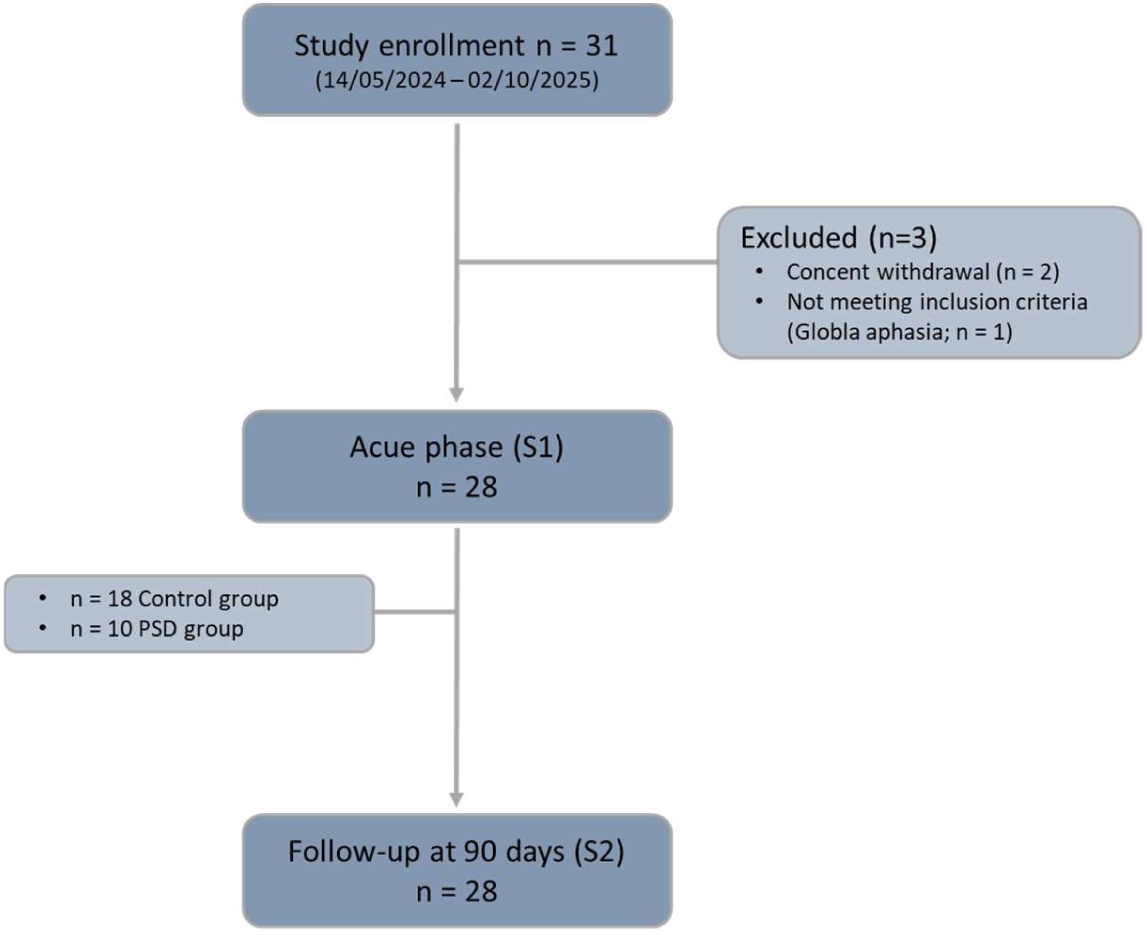
Flowchart of patients. Flow diagram presenting the enrolment of 31 patients. Before the first assessment three patients has been excluded due to consent withdrawal and global aphasia following clinical deterioration. After the first assessment 10 patients developed PSD and 18 stroke patients did not. Abbreviations: PSD = post-stroke delirium. S1 = Session 1, S2 = session 2).

Demographic and clinical characteristics including age, sex, education, pre-stroke cognitive and functional status, stroke location and hemisphere as well as acute reperfusion therapies, did not significantly differ between groups (Table 2). However, patients who subsequently developed PSD had higher NIHSS scores on admission than those without PSD (median, 9 [IQR, 13.75] versus 2.5 [IQR, 5.25]; *U* = 41.000, *p* = 0.018).

**Table 2.** Demographics and Clinical Characteristics.

|  | **All**<br>**n=28** | **PSD**<br>**n=10** | **Control**<br>**n=18** | **p-value** |
| --- | --- | --- | --- | --- |
| Age, mean years (SD) | 77 (9.6) | 80 (7.8) | 75(10.2) | 0.18 ° |
| Sex, n (%) |  |  |  | 0.43 + |
| Male | 15 (54) | 4 (40) | 11 (61) |  |
| Female | 13 (46) | 6 (60) | 7 (39) |  |
| Education, mean years (SD) | 11.7 (3.6) | 10.7 (3.6) | 12.2(3.6) | 0.24 + |
| Cognitive level prior stroke (CFQ),<br>mean (SD)<br>missing values n=4 (all in PSD group) | 27.2 (11.4) | 27.8 (12.2) | 26.9 (11.5) | 0.87° |
| Stroke location, n (%) |  |  |  | 0.28 * |
| Anterior circulation | 25 (89) | 8 (80) | 17 (94) |  |
| Posterior circulation | 3 (11) | 2 (20) | 1 (6) |  |
| Stroke hemisphere, n (%) |  |  |  | 0.80 * |
| Left | 13 (46) | 4 (40) | 9 (50) |  |
| Right | 14 (50) | 6 (60) | 8 (44) |  |
| Bilateral | 1 (4) | (0) | 1 (6) |  |
| pmRS, median (IQR) | 0 (1) | 0.5 (1) | 0 (0) | 0.160 # |
| NIHSS on admission, median (IQR) | 5 (8) | 9 (13.75) | 2.5 (5.25) | **0.018** # |
| Acute Therapy |  |  |  |  |
| IVT, n (%) | 10 (36) | 3 (30) | 7(39) | 0.70 + |
| Thrombectomy, n (%) | 6 (21) | 2 (20) | 4 (22) | 1.00 + |

Abbreviations: CFQ = Cognitive Failure Questionnaire, IQR = Interquartile Range, IVT = intravenous thrombolysis, NIHSS = National Institute of Health Stroke Scale, PSD = post-stroke delirium, pmRS = premorbid modified Rankin Scale, SD = Standard deviation. Statistical tests: # = Mann-Whitney-U-Test. \* = Chi-Square-Test, + = Fisher's-exact-test, ° = T-Test for independent sample.

### 3.2. Primary outcome

Stroke patients who developed PSD had a significantly worse global cognitive outcome with more severe gPSCI (*U* = 19.500, *p* < 0.001; Table 3, Figure 3) than stroke patients without subsequent PSD, at 90 days post-stroke.

**Figure 3.**
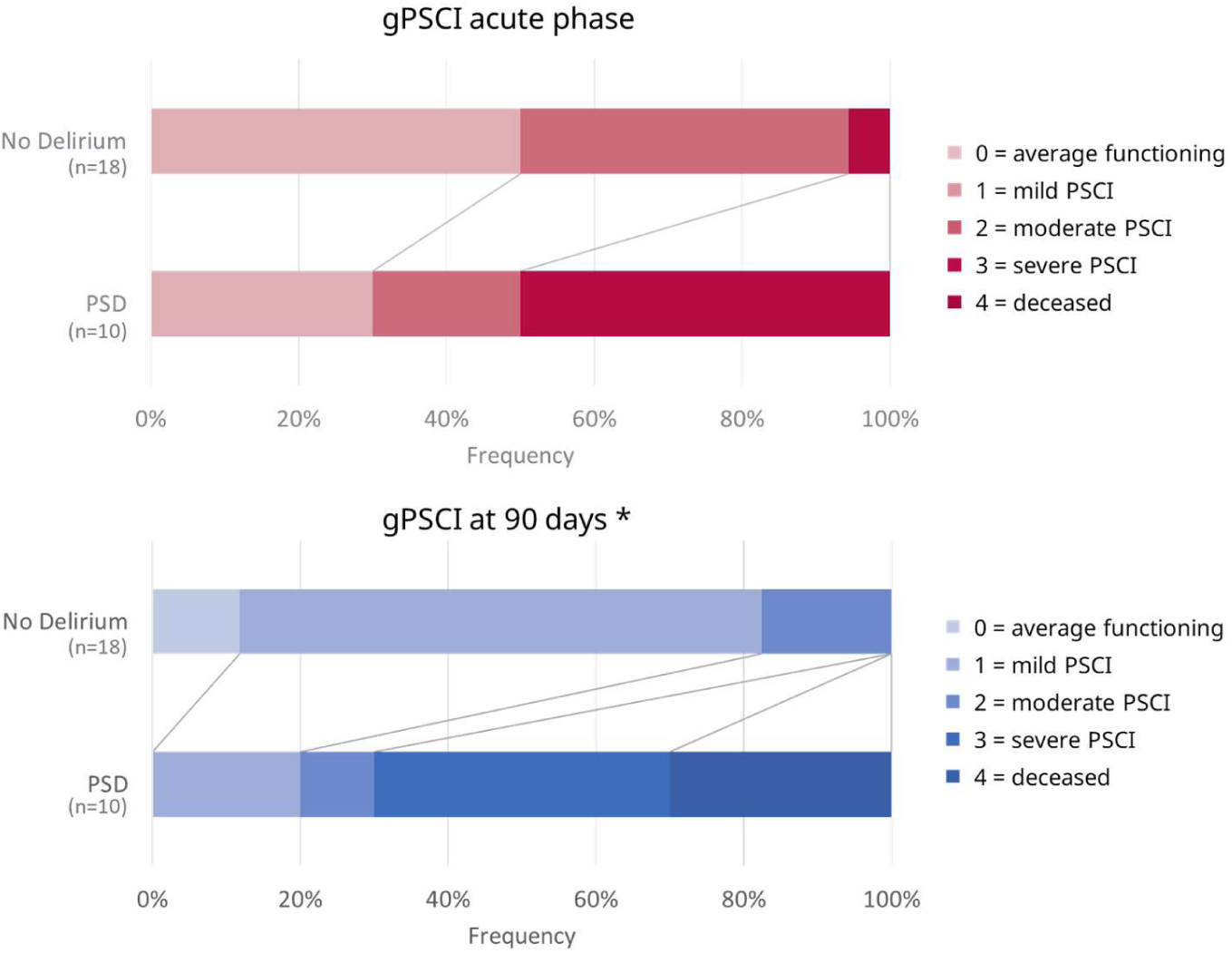
Distribution of global post-stroke cognitive impairment (gPSCI) during the acute phase, prior PSD onset and at 90 days post-stroke. gPSCI scores range from 0 to 4, with highest scores indicating increased cognitive impairment, and 4 indicating death. Asterisk indicates significant group differences. Abbreviations: gPSCI = global post-stroke cognitive impairment, PSD = Post-stroke delirium.

**Table 3:** Clinical course and outcomes at during the acute phase and 90 days post-stroke.

| Outcomes | All<br>n=28 | PSD<br>n=10 | Control<br>n=18 | p-value |
| --- | --- | --- | --- | --- |
| gPSCI during acute phase (prior PSD), median<br>(IQR) | 2 (2) | 3 (2) | 2 (1) | 0.057 # |
| dsPSCI attention & processing speed during acute phase (prior PSD), median (IQR) | 1 (1.75) | 2.5 (2) | 1 (1) | **0.003**<sup>#</sup> |
| dsPSCI executive functions during acute phase (prior PSD), median (IQR) | 3 (2) | 3 (2) | 3 (2) | 0.80<sup>#</sup> |
| dsPSCI verbal memory during acute phase (prior PSD), median (IQR) | 1 (2) | 1.5 (2.25) | 1 (1.25) | 0.099<sup>#</sup> |
| dsPSCI language during acute phase (prior PSD), median (IQR) | 1 (2.75) | 2.5 (3) | 1 (2) | 0.146<sup>#</sup> |
| dsPSCI visuospatial functions during acute phase (prior PSD), median (IQR) | 0 (2.75) | 3 (2.25) | 0 (1) | **0.004**<sup>#</sup> |
| gPSCI at 90d, median (IQR) | 1(1.75) | 3(2.25) | 1(1) | **< 0.001**<sup>#</sup> |
| dsPSCI attention & processing speed at 90d, median (IQR) | 1 (1.75) | 3 (3) | 0 (1) | **< 0.001**<sup>#</sup> |
| dsPSCI executive functions at 90d, median (IQR) | 3 (2) | 3 (1) | 2.5 (3) | **0.012**<sup>#</sup> |
| dsPSCI verbal memory at 90d, median (IQR) | 0 (2) | 1 (3) | 0 (1) | 0.23<sup>#</sup> |
| dsPSCI language at 90d, median (IQR) | 1 (2) | 2.5 (3.25) | 1 (2) | **0.031**<sup>#</sup> |
| dsPSCI visuospatial functions at 90d, median (IQR) | 0 (1) | 0.5 (4) | 0 (1) | 0.175<sup>#</sup> |
| Cognitive Recovery at 90d, n (%) | 12 (43) | 1 (10) | 11 (61) | **0.016**<sup>+</sup> |
| NIHSS at 90d, median (IQR)<br>missing values n=3 (diseased; all in PSD group) | 1 (2.5) | 2 (4) | 1 (2) | 0.22<sup>#</sup> |
| mRS at 90d, median (IQR) | 3 (2.75) | 4.5 (3) | 2 (2) | **0.004<sup>#</sup>** |
| Good Outcome (mRS 0-2) at 90d, n (%) | 11 (39) | 1 (10) | 10 (56) | **0.041<sup>+</sup>** |
| Favorable Outcome (mRS ≤ 3) at 90d, n (%) | 18 (64) | 3 (30) | 15 (83) | **0.013<sup>+</sup>** |
| Mortality at 90d, n (%) | 3 (11) | 3 (30) | 0 (0) | **0.041<sup>+</sup>** |
| Post-stroke Depression at 90d, n (%)<br>missing values n=3 (diseased; all in PSD group) | 13 (52) | 5(71) | 8(44) | 0.37<sup>+</sup> |
| Quality of life (Mental) at 90d, mean (SD) | 45.5 (13.3) | 38.9 (15.9) | 49.5 (10.3) | **0.021<sup>°</sup>** |
| Quality of life (Physical) at 90d, mean (SD) | 34.9 (13.4) | 30.1 (12.4) | 37.7 (13.5) | 0.156<sup>°</sup> |

Abbreviations: 90d = 90 days post-stroke follow-up, gPSCI = global Post-Stroke Cognitive Impairment, IQR = Interquartile Range, mRS = modified Rankin Scale, NIHSS = National Institute of Health Stroke Scale, PSD = Post Stroke Delirium, S1 = session 1 during acute state and prior PSD. Statistical tests: # = Mann-Whitney-U-Test. \* = Chi-Square-Test, + = Fisher's-exact-test, ° = T-Test for independent sample. Each variable is displayed with median and interquartile range (IQR) or counts and percentages, where applicable.

#### Secondary outcomes

During the acute phase and before PSD onset, global PSCI did not significantly differ between groups (*U* = 50.500, *p* = 0.057; Table 3, Figure 3). Domain-specific PSCI analyses revealed distinct temporal patterns of cognitive impairment. During the acute phase, patients who subsequently developed PSD showed greater impairment in attention and processing speed (U= 29.500; *p* = 0.003) and visuospatial-construction (U= 31.500; *p* = 0.004; Table 3, Figure 4, Figure S1). At 90 days post-stroke, the group-difference in attention and processing speed remained significant (*U* = 21.000, *p* < 0.001), with additional significant group differences in the domains executive functions (*U* = 38.500, *p* = 0.012) and language (*U* = 45.500, *p* = 0.031), whereas verbal memory (*U* = 64.000, *p* = 0.226) and visuospatial-construction (*U* = 61.000, *p* = 0.175) did not significantly differ between groups (Table 3, Figure 5, Figure S1).

**Figure 4.**
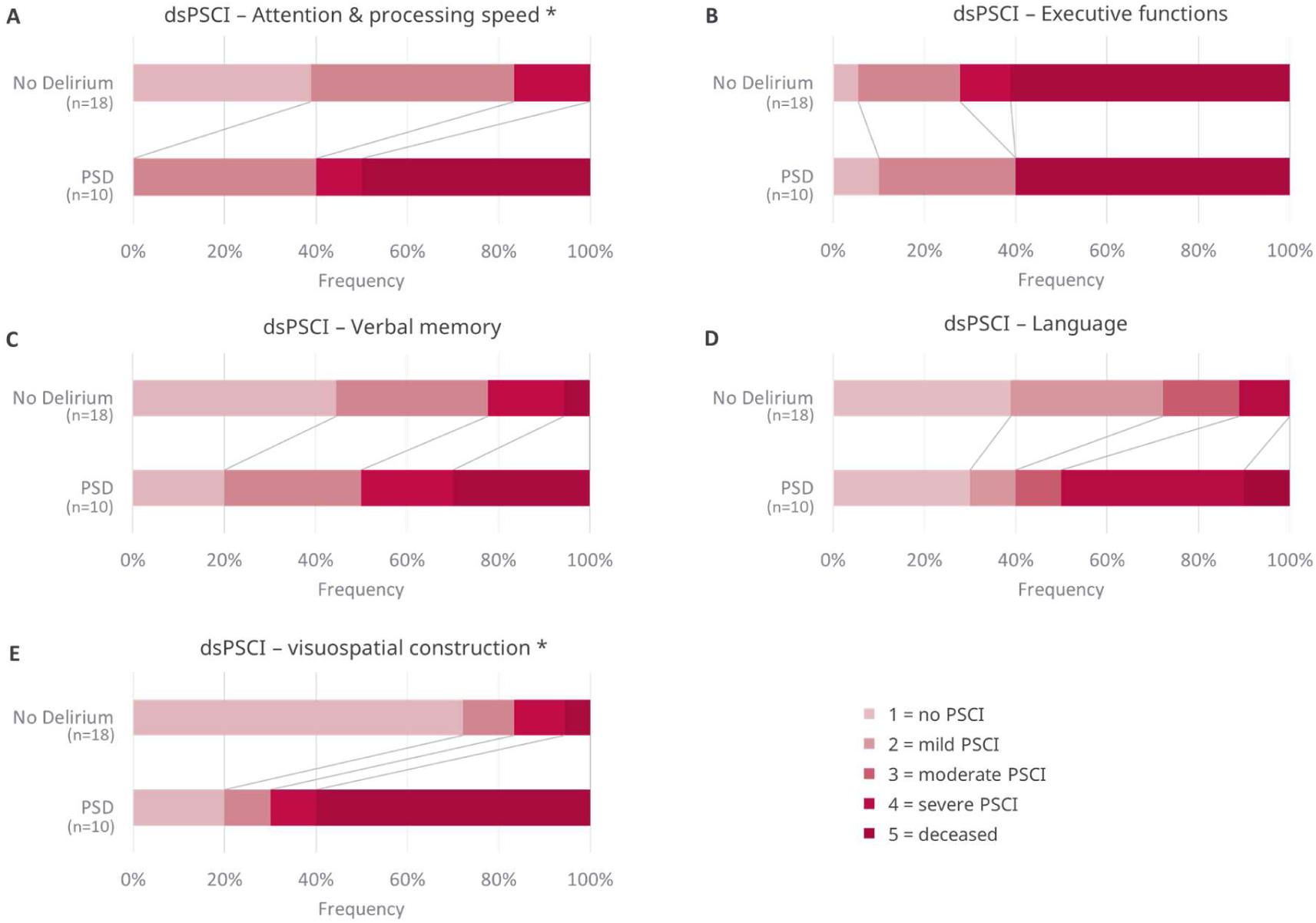
Distribution of domain-specific post-stroke cognitive impairment (dsPSCI) during the acute-phase and before PSD onset. dsPSCI scores range from 0 to 4, with highest scores indicating increased cognitive impairment, and 4 indicating death. Asterisk indicates significant group differences. Abbreviations: ds = domain specific post-stroke cognitive impairment, PSD = Post-stroke delirium.

**Figure 5.**
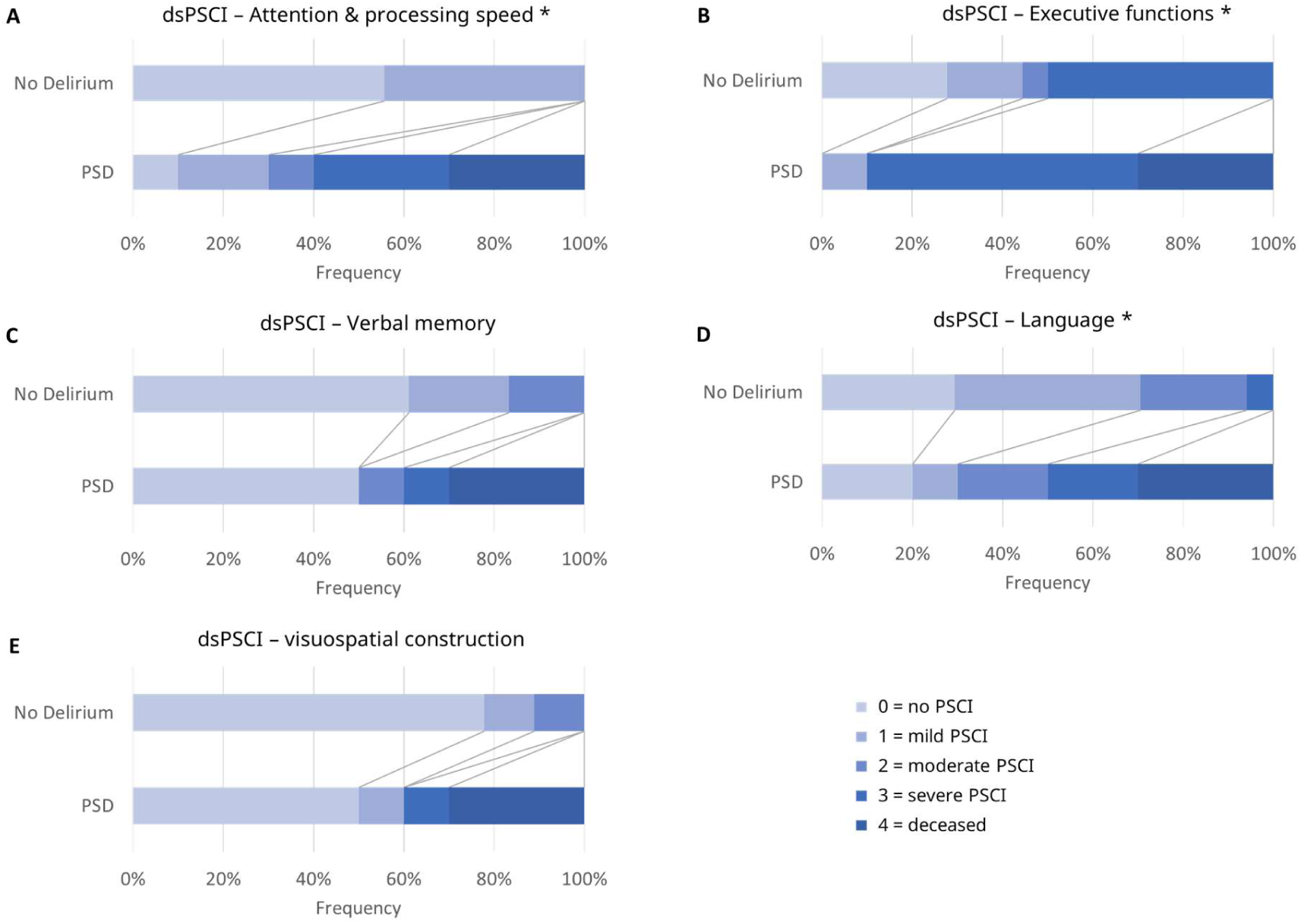
Distribution of domain-specific post-stroke cognitive impairment (dsPSCI) at 90 days post-stroke. dsPSCI scores range from 0 to 4, with highest scores indicating increased cognitive impairment, and 4 indicating death. Asterisk indicates significant group differences. Abbreviations: ds = domain specific post-stroke cognitive impairment, PSD = Post-stroke delirium.

Further, at 90 days post-stroke, patients with PSD showed a significantly worse functional outcome (mRS; *U* = 31.500, *p* = 0.004, Table 3, Figure 6), higher mortality rate (*p* = 0.037), lower rates of cognitive recovery (*p* = 0.016) and a lower mental quality of life (*p* = 0.021) at 90 days (Table 3). In an exploratory ordinal logistic regression, PSD was associated with higher odds of a worse mRS category (*OR*, 14.81 [95% *CI*, 2.69–81.62]; *p* = 0.002). NIHSS at 90 days did not significantly differ between groups (*p* = 0.220, Table 3).

**Figure 6.**
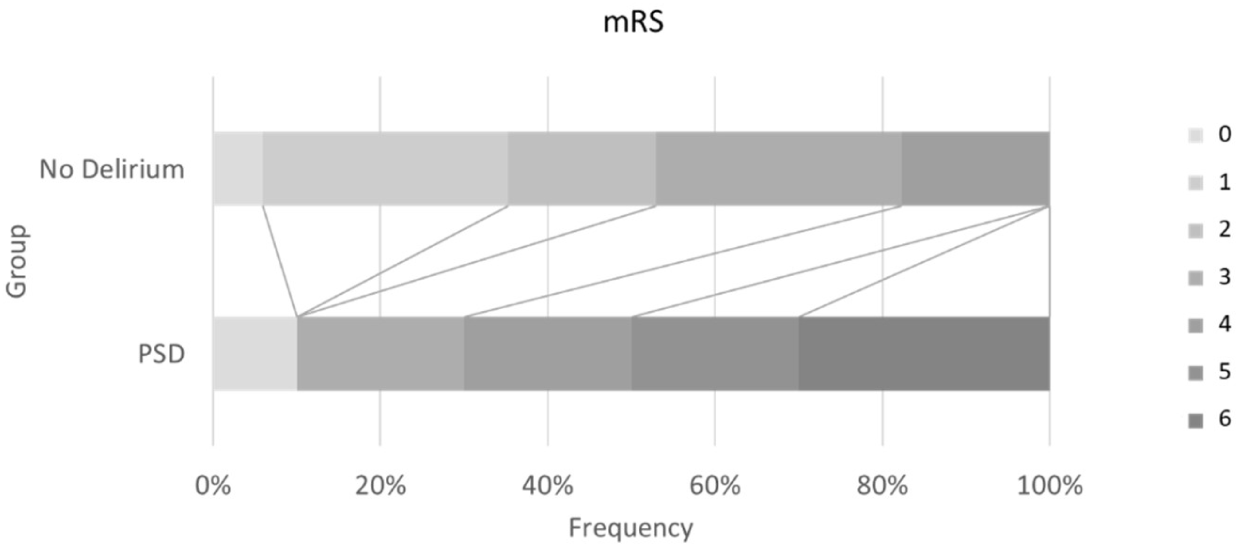
Distribution of modified Rankin Scale (mRS) at 90 days post-stroke. mRS scores range from 0 to 6, with highest scores indicating increased disability, and 6 indicating death.

In a random forest classification analysis of kinematic key features derived from the figure copying task, performed consistently above the baseline chance level of 66.66%. Across the repeated iterations, mean classification accuracies were 81.50% (SE = 0.14%) for the control model using gPSCI, 78.89% (SE = 0.15%) for the drawing model, and 75.19% (SE = 0.16%) for the combined model (Figure 7). To generate final, stable model predictions and account for algorithmic variance, a majority-vote consensus was applied across the repetitions for each patient. Following this aggregation, both the control and the drawing model achieved an identical consensus accuracy of 81.50% (SE = 1.50%), while the combined model reached 77.78% (SE = 1.60%). An omnibus Cochran’s Q test confirmed that there were no statistically significant differences in overall predictive performance across the three feature sets (Q(2) = 0.25, p = 0.882), establishing the non-inferiority of the drawing features to the clinical baseline.

**Figure 7.**
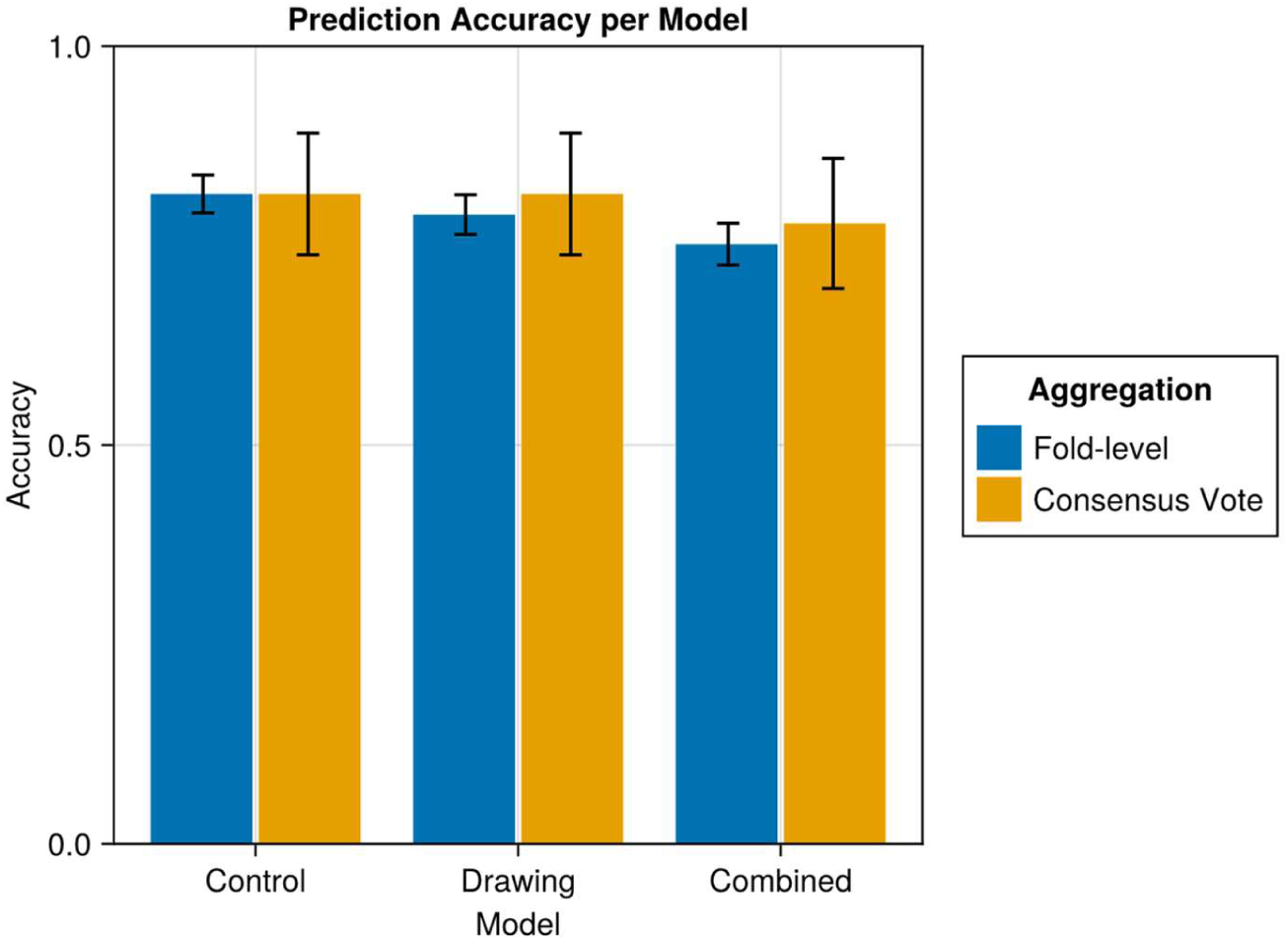
Prediction accuracy of the control (using global PSCI), drawing, and combined models using fold-level and majority-vote consensus aggregation. Bars represent mean classification accuracy, and error bars indicate the standard error (SE).

## 4. Discussion

This longitudinal study characterized global and domain-specific PSCI in stroke patients with and without subsequent PSD using tablet-based multidomain neuropsychological assessment during the acute phase and 90 days post-stroke. Here we present a study, where cognitive assessment was performed during the early acute phase post-stroke (within 7hrs after stroke) compared with most of the existing literature, where cognition has been mostly assessed in early chronical state or chronical state ^18–20^. Furthermore, this is one of the few studies, to assess cognitive function in stroke patients before the onset of PSD, providing the opportunity to assess subsequent cognitive trajectories related to PSD and identify early cognitive vulnerability and patients at risk of PSD development.

First, we demonstrate that PSD is significantly associated with a more severe global PSCI and significantly lower rates of cognitive recovery. This finding is consistent with previous studies of PSD that assessed global cognition using sum scores or cut-off values, for example, the MoCA score at 90 days post-stroke ^16, 17, 56^.

Second, PSD is associated with a distinct temporal pattern of cognitive impairment. During the acute phase and prior PSD onset, no significant between-group difference was observed for global PSCI but further analyses showed that, patients who subsequently developed PSD, already show significantly more severe impairment in the domains of attention and processing and visuospatial construction. At 90 days post-stroke, the group-difference in attention and processing speed remained significant and additional group-differences were demonstrated in the domains of executive functions and language compared with stroke patients without PSD. In general, previous findings have shown that long-term PSCI, irrespective of PSD, often persists in domains such as attention, processing speed or visuo-executive functions ^16, 54, 55^. A previous small-sample size retrospective study on PSD showed that the cognitive domains most affected in patients with previous delirium were memory, language, visual construction and executive functioning^57^. In line with these findings, our prospective longitudinal study demonstrates an additional effect of PSD on specific cognitive domains. One possible explanation might be that persistent attentional deficits associated with PSD may impair higher-order cognitive processes such as visuo-executive functions or language, which require intact attentional resources ^58^. Thus, recognizing PSD as a distinct clinical condition characterized by specific patterns of cognitive impairment during the early post-stroke phase, may help clinicians identify patients at risk and potentially facilitate earlier preventive interventions.

Third, in an exploratory analysis, acute-phase task-derived kinematic parameters (obtained before PSD onset) showed discriminatory ability between patients who subsequently developed PSD and those who did not. These findings suggest that kinematic parameters derived from a single figure-copying task may capture information relevant to PSD distinct from conventional cognitive scores. Figure copying engages multiple higher-order cognitive processes, including attention and processing speed, visuospatial-constructional abilities, and executive functioning, integrating information across several cognitive domains within a single task. The comparable discriminatory performance of selected digital parameters and established cognitive measures highlights the potential utility of app-based kinematic assessment in acute stroke settings. However, the small sample size presents a risk of the models fitting to sample-specific artifacts, which precludes firm conclusions regarding absolute predictive performance. Validation in larger, independent cohorts is required to establish generalizability.

Further, our results demonstrate that patients with PSD showed a significantly worse functional outcome (mRS), significantly lower rates of favorable (mRS < 3) or good (mRS < 2) outcome, as well as significantly higher mortality rates, which is in line with previous research ^1, 6–10, 12, 13^ and underscore the urgent need for suitable diagnostic and prognostic measures ^11^. Despite major advances in stroke care and treatment over the last decades, mortality associated with PSD has remained high ^2, 9^.

Furthermore, patients with PSD showed significantly lower mental quality of life compared to non-delirious stroke patients. There was no significant effect on physical quality of life despite the significant difference in functional outcomes as measured by the mRS. Most previous studies regarding quality of life ^1, 6–13^ have reported lower quality of life in patients with PSD in general. One possible explanation for the differences between our findings and previous studies is that PSCI in general has a substantial impact on quality of life ^23^, while the assessment of quality of life varies depending on the questionnaires used, with some not providing separate mental and physical components. ^59^. Future studies should investigate the differential burden of PSD on mental and physical quality of life more closely.

Another important aspect to consider is the method used to assess PSCI. The approach used in this study highlights the importance of a precise and comprehensive assessment of cognitive function across multiple cognitive domains. Unfortunately, only few longitudinal studies have investigated and characterized domain-specific cognitive trajectories after stroke ^55^ ^60, 61^. In many studies, the primary outcome relies on a dichotomous classification of PSCI based on a single cutoff threshold or on sum scores derived from cognitive screening batteries, such as the MoCA or MMSE ^13, 17, 19, 21^. However, these approaches may result in substantial information loss, as they do not capture the full spectrum of cognitive impairments across individual domains. Furthermore, dichotomous or sum-score-based approaches have been reported to be not sensitive enough to detect subtle PSCI ^12, 14, 62^ and may not adequately account for differential dropout due to mortality during follow-up. In particular, reliance on a total score from a paper-based screening assessment may obscure impairments in specific cognitive domains that are detectable with a more comprehensive multidomain assessment. Therefore, future longitudinal studies should account for this potential bias and consider outcome measures and analytical approaches that appropriately incorporate mortality during follow-up.

This study has several methodological strengths. First, its prospective longitudinal design enabled cognitive assessment during the acute phase after stroke, within 48 hours of stroke onset, before the occurrence of PSD and follow-up 90 days post-stroke. This temporal framework addresses an important limitation of previous studies, which have predominantly characterized cognition after the acute phase or following the onset of delirium and provides an opportunity to assess subsequent cognitive trajectories in relation to the development of PSD.

Second, the very low dropout rate demonstrates the feasibility of repeated multidomain cognitive assessment during the acute stroke phase despite the clinical challenges of early post-stroke assessment.

Third, comprehensive neuropsychological assessment across multiple cognitive domains, complemented by tablet-based digital parameters, enabled standardized and quantitative characterization of domain-specific cognitive profiles. The observed ability of acute-phase cognitive measures and selected kinematic parameters to classify patients according to 90-day PSD status suggests potential prognostic relevance and support the potential scalability of computationally derived features for early risk stratification. The use of continuous measures of global and domain-specific cognitive performance further preserved information across the spectrum of impairment and may provide greater sensitivity for characterizing heterogeneous cognitive trajectories. These methodological features provide a basis for prospective multicenter studies to validate early cognitive markers and establish their prognostic value.

Finally, matching and balancing of the PSD and stroke control groups for clinically relevant factors, including lesion location and acute stroke treatment, strengthened between-group comparability and reduced potential confounding. Although residual confounding cannot be excluded, these features strengthen the internal validity of the observed associations.

This study has several limitations. First, the small sample size limits statistical power and precluded robust adjustment for potential confounders. In particular, patients who developed PSD had higher admission NIHSS scores, and stroke severity is a well-established risk factor for both PSD and adverse cognitive outcomes. Thus, residual confounding by stroke severity cannot be excluded. Second, the exclusion of patients with global aphasia limits generalizability to the broader stroke population. Finally, the exploratory analyses of digital cognitive parameters should be interpreted cautiously and viewed as a proof of principle approach, given the small sample size and require validation in independent cohorts.

Despite these limitations, our findings suggest that PSD is associated with a distinct cognitive trajectory, with early domain-specific deficits preceding more pronounced global and domain-specific cognitive impairment at 90 days. Larger multicenter longitudinal studies are needed to validate these findings and determine the clinical utility of early multidomain and digital cognitive assessment.

## Data Availability

The datasets generated during the current study are not publicly available due General Data Protection Regulation restrictions but are available from the corresponding author on reasonable request.

## Acknowledgements

We thank Maria Friederich for her assistance in conducting this study.

## Authors’ contributions

L.S.G and A.M. designed the study protocol, provided the original idea of the work. L.S.G., A.M. and F.W. drafted the manuscript. L.S.G., Se.K. acquired data with the help of A.J., So.K. and C.S. and J.S.P. L.S.G., F.W., Se.K., and C.M. performed the analysis under the supervision of A.M. and U.Z.. C.M., C.G., and F.W. developed and programmed the app-based assessment. All authors substantively revised and approved the final version of this manuscript.

Abbreviations

CFQ: Cognitive failures questionnaire
CI: Confidence interval
dsPSCI: domain-specific post-stroke cognitive impairment
90d: Follow-up at 90-days post-stroke
GCS: Glasgow Coma Scale
GDS: Geriatric Depression Scale
gPSCI: global post-stroke cognitive impairment
ICDSC: Intensive Care Delirium Screening Checklist
ICU: Intensive Care Unit
IQCODE: Informant Questionnaire on Cognitive Decline in the Elderly
IQR: Interquartile Range
IVT: Intravenous Thrombolysis
LOOCV: Leave-one-out cross-validation
MMSE: Mini-Mental-State Examination
MoCA: Montreal Cognitive Assessment Test
mRS: modified Rankin Scale
NAB: Neuropsychological assessment battery
NIHSS: National Institutes of Health Stroke Scale
OR: Odds ratio
pmRS: pre-modified Rankin Score (before hospital admission)
PSD: Post-stroke delirium
PSCI: Post-stroke cognitive impairment
TMT: Trail making test
TuCAN: Tuebingen Cognitive Assessment for Neuropsychiatric Disorders
SE: Standard Error
SF-12: Short-Form Health Survey (12 items)
S1 – S2: Session 1 – Session 2
VASCOG: International Society for Vascular Behavioral and Cognitive Disorders
WM: Working memory

